# Proteomic biomarker candidates inversely associated with menopausal symptoms in midlife women

**DOI:** 10.64898/2026.08.17.26360645

**Authors:** Maki Sasanuma, Misa Kuroki, Hirotaka Tabata, Atsushi Kajiwara, Akiko Shiraki, Rehab F. Abdelhamid, Yukoh Nakazaki, Miho Takao

## Abstract

**Objectives:** Menopausal symptoms are heterogeneous and commonly assessed by questionnaires. We explored serum two-dimensional gel electrophoresis (2-DE) protein spots associated with menopausal symptom burden.

**Methods:** This exploratory cross-sectional study included 27 women aged 45-55 years. A total of 550 matched serum 2-DE spots were quantified. A frequency-adjusted symptom burden score was calculated as the sum of severity × frequency products across 10 symptoms. Spots were screened using Spearman rank correlation with Benjamini-Hochberg false discovery rate (FDR) adjustment, followed by qualitative image review. Spots #285 and #636 were prioritized for vasomotor and psychological domain analyses.

**Results:** The median age was 51.0 years; 13 participants were menstruating and 14 were amenorrheic. The median overall symptom burden score was 45.0 (interquartile range, 6.5- 58.5) and was inversely correlated with spots #285 and #636. Spot #285 was inversely correlated with vasomotor symptom score, including inverse correlations in both menstrual- status groups. Spot #636 was inversely correlated with psychological symptom score overall, with a stronger descriptive correlation among menstruating participants. Neither candidate remained significant after FDR adjustment.

**Conclusions:** Spots #285 and #636 are hypothesis-generating candidates requiring molecular identification, analytical validation, multiplicity-aware confirmation, and independent replication.

## 1. Introduction

The menopausal transition is accompanied by diverse symptoms, including hot flushes, sweating, sleep disturbance, irritability, depressed mood, fatigue, and musculoskeletal complaints. Symptom type and severity vary substantially among individuals, and clinical assessment therefore relies heavily on patient-reported information and structured questionnaires [1]. Serum estradiol (E2) and follicle-stimulating hormone (FSH) are useful indicators of reproductive endocrine status [2]. However, their fluctuating secretion patterns and limited correspondence with individual symptom profiles may restrict their ability to objectively characterize symptom burden [3]. An objective biological measure that complements subjective symptom assessment could support symptom stratification, longitudinal monitoring, and treatment decisions.

Vasomotor symptoms are among the hallmark manifestations of the menopausal transition and can persist for several years, substantially affecting quality of life [4]. Psychological symptoms and mental health are also clinically important during this period. Hormonal fluctuations during the menopausal transition have been linked to complex and potentially bidirectional changes in mental health, although such symptoms are not experienced by all women [5,6]. The present study therefore focused particularly on vasomotor and psychological symptom burden.

Proteins are direct functional components of biological processes and can reflect changes in abundance, processing, and post-translational state. Recent reviews have emphasized the potential value of proteomic research in menopausal health [7]. Proteins associated with cardiovascular disease and osteoporosis in peri- and postmenopausal women have been identified [8–12], and proteomic changes linked to dietary interventions and hormone replacement therapy have also been reported [13–16]. These findings suggest that proteomic approaches may contribute to improved biological characterization of menopausal health. In the present study, we used two-dimensional gel electrophoresis (2-DE) for proteomic profiling. This method separates serum proteins according to isoelectric point and molecular mass, enabling broad visual comparison of protein spot patterns without restricting the analysis to a predefined analyte panel. Serum 2-DE profiling has previously been applied in exploratory biomarker studies of sepsis and complications following hematopoietic stem-cell transplantation [17,18].

The present pilot study explored associations between serum 2-DE protein spot intensities and menopausal symptom burden using symptom scores as continuous variables. We first screened 550 protein spots for associations with overall frequency-adjusted symptom burden and then reviewed candidate spots qualitatively on the 2-DE images. Spots #285 and #636 were prioritized on the basis of statistical screening and analytical suitability and were subsequently examined in relation to vasomotor and psychological symptom domains, respectively. Correlations for the two candidates were also estimated separately in menstruating and amenorrheic participants. The analytical workflow is shown in Fig. 1A.

**Fig. 1.**
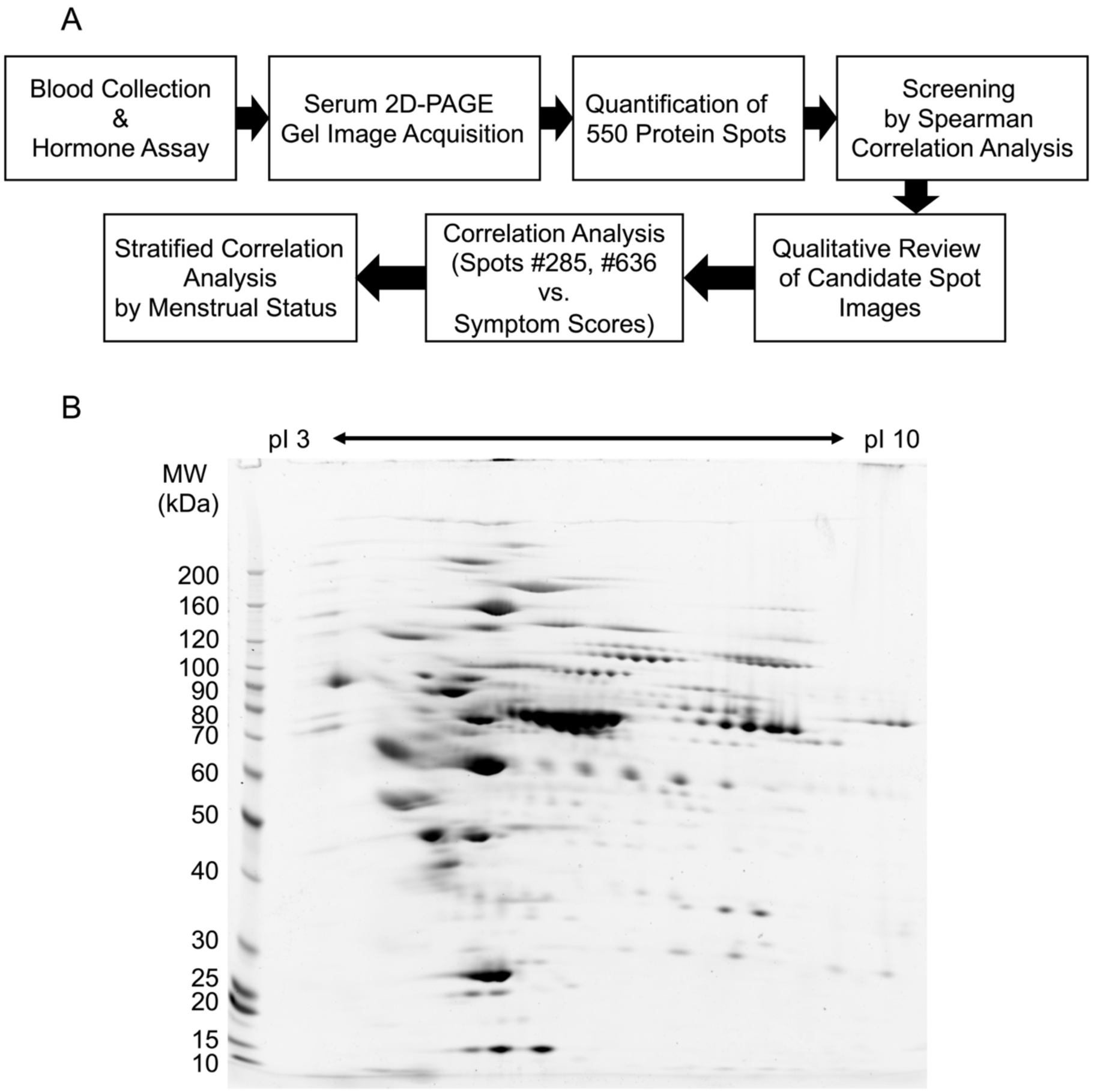
Overview of the analytical workflow and representative serum 2-DE protein profile. (A) Schematic overview of the analytical workflow. Blood samples were collected for hormone measurements, and serum proteins were separated by two-dimensional gel electrophoresis (2-DE). A total of 550 protein spots were quantified and screened for associations with the overall frequency-adjusted symptom burden score using Spearman rank correlation. Candidate spots were subsequently reviewed qualitatively on the 2-DE images, and spots #285 and #636 were prioritized for domain-specific analyses. Correlations for the selected candidate spots were then estimated separately in menstruating and amenorrheic participants. (B) Representative 2-DE gel image of serum proteins showing the overall distribution of quantified protein spots. Proteins were separated according to isoelectric point (pI 3–10) in the first dimension and molecular mass in the second dimension.

## 2. Materials and Methods

### 2.1 Study design and participants

This was an exploratory cross-sectional study of women aged 45-55 years. Twenty-nine participants were available in the study dataset. Two participants (MT11 and MT20) were excluded from the analysis because their self-reported menstrual status was inconsistent with the measured E2 and FSH values, leaving 27 participants in the analytic cohort. Participants were publicly recruited paid volunteers and were required to meet the following inclusion criteria:

1. Individuals who received a full explanation of the study purpose and procedures, had the capacity to provide consent, understood the study, volunteered to participate, and provided informed consent.
2. Japanese women aged 45-55 years at the time of consent.
3. Individuals experiencing menopausal symptoms.
4. For menstruating individuals, a regular menstrual cycle of 25 to 38 days.

The exclusion criteria were as follows:

1. Individuals receiving pharmacological treatment for chronic disease or with a history of severe illness.
2. Individuals routinely using pharmaceuticals or health food products (such as Foods for Specified Health Uses, Foods with Function Claims, and Foods with Nutrient Function Claims) that could affect menopausal symptoms.
3. Individuals who had participated in another research study within 1 month before the start of this study or were scheduled to participate in another research study after providing consent.
4. Individuals deemed ineligible by the study supervisor or principal investigator.
5. Individuals who were lactating, pregnant, or planning or wishing to become pregnant during the study period.

Menstrual status was classified according to the response obtained at blood collection. Participants who reported being menopausal or amenorrheic were assigned to the amenorrheic group; all remaining participants were assigned to the menstruating group.

### 2.2 Ethics

The study protocol was reviewed and approved by the Ethics Committee of Healthcare Systems Co., Ltd. (approval no. 2426; October 31, 2024). The study was conducted in accordance with the ethical principles of the Declaration of Helsinki and the Ethical Guidelines for Medical and Biological Research Involving Human Subjects issued by the Ministry of Education, Culture, Sports, Science and Technology; the Ministry of Health, Labour and Welfare; and the Ministry of Economy, Trade and Industry (2021). All participants received an explanation of the study purpose, procedures, potential risks and benefits, the voluntary nature of participation, and their right to withdraw without disadvantage. Written informed consent was obtained from all participants before participation in the study.

### 2.3 Symptom assessment

Participants reported the severity and frequency of 10 symptoms corresponding to items used in the Simplified Menopausal Index (SMI): hot flushes, sweating, coldness in the lower back and extremities, shortness of breath/palpitations, sleep disturbance, irritability, depressed mood, headache/dizziness/nausea, fatigue, and shoulder stiffness/lower-back pain/limb pain [19,20]. For the present analyses, symptom severity was scored as 0 (none), 1 (mild), 2 (moderate), or 3 (severe). Frequency was scored as 1 (approximately monthly), 2 (several times per month), 3 (approximately weekly), 4 (approximately three times per week), or 5 (almost daily); symptoms rated as having no severity were assigned a score of 0. Each symptom score was calculated as severity × frequency. The overall frequency-adjusted symptom burden score was calculated as the sum of the 10 symptom scores (range, 0-150). The vasomotor symptom score was calculated as the sum of the hot-flush and sweating scores (range, 0-30), and the psychological symptom score as the sum of the depressed mood, irritability, and sleep disturbance scores (range, 0-45).

### 2.4 Blood collection and hormone measurements

Blood was collected and serum was prepared for E2, FSH, and proteomic analyses. Blood collection was performed by Healthcare Systems Co., Ltd. (Aichi, Japan), and E2 and FSH measurements were performed by Fukuyama Clinical Laboratory Center (Hiroshima, Japan). The lower limit of quantification for E2 was 5.0 pg/mL. Values below this limit were reported as <5.0 pg/mL and were provisionally assigned a value of 5.0 pg/mL for calculation of descriptive medians and quartiles.

### 2.5 Two-dimensional gel electrophoresis and image analysis

To deplete abundant proteins, serum samples were preprocessed using the High Select Top14 Abundant Protein Depletion Kit (Cat. A36372; Thermo Fisher Scientific, MA, USA). The samples were purified using the PAGE Clean Up Kit (Cat. 06441-50; Nacalai Tesque, Kyoto, Japan), and proteins were dissolved in DeStreak Rehydration Solution (Cat. 17600319; Cytiva, MA, USA). Total protein concentration was measured using the 2-D Quant Kit (Cat. 80648356; Cytiva), and 15 µg of total protein from each sample was subjected to 2-DE. In the first dimension, proteins were separated according to isoelectric point (pH 3-10); in the second dimension, they were separated according to molecular mass. Two-dimensional gel electrophoresis was performed as described by Wong et al. [18]. Proteins were stained with SYPRO™ Protein Gel Stain (Cat. S21900; Thermo Fisher Scientific), and gel images were acquired using a FluoroPhoreStar 3000 system (Cat. 3000-22; Anatech, Tokyo, Japan). Protein spots were detected using SameSpots image-analysis software (version 5.1.012; TotalLab, Gosforth, UK), and spot intensities were quantified and normalized to the total spot intensity in each gel image.

### 2.6 Statistical analysis

Continuous participant characteristics are summarized as medians and interquartile ranges (IQRs), and categorical variables as counts and percentages. Because normalized spot volumes were skewed and the sample size was limited, nonparametric methods were used for the principal analyses. The principal exploratory screening analysis evaluated the association between the frequency-adjusted symptom burden score and each of the 550 protein spots.

Associations between normalized spot volumes and symptom scores were assessed using two-sided Spearman rank correlation coefficients. For the overall frequency-adjusted symptom burden score, P values from the 550 spot-wise tests were adjusted using the Benjamini-Hochberg false discovery rate (FDR) procedure. Spots with an absolute Spearman correlation coefficient of at least 0.50 and a nominal P value < 0.01 were retained as statistical candidates. These candidates were then qualitatively reviewed on the 2-DE images for spot definition, signal intensity, separation from neighboring spots, and matching consistency across gels. Spots #285 and #636 were prioritized on the basis of the combined statistical and image review for subsequent domain-specific analyses. Secondary analyses examined the association between spot #285 normalized volume and vasomotor symptom score and between spot #636 normalized volume and psychological symptom score. Domain-specific P values were nominal and were not adjusted for the candidate-selection process. Correlations were also estimated separately in menstruating and amenorrheic participants. Spearman ρ values are reported with percentile 95% confidence intervals. A two-sided P < 0.05 was used as a descriptive threshold. Analyses were performed using GraphPad Prism version 11.0.2.

## 3. Results

### 3.1 Participant characteristics

Twenty-seven participants were included in the analysis: 13 were classified as menstruating and 14 as amenorrheic. The median age was 51.0 years (IQR, 49.0-53.5 years), median BMI was 21.0 kg/m² (IQR, 19.4-22.8 kg/m²), median FSH was 44.8 mIU/mL (IQR, 10.3-70.2 mIU/mL), and median E2 was 45.6 pg/mL (IQR, 5.0-105.5 pg/mL). Ten participants had E2 values below 5.0 pg/mL, all of whom were in the amenorrheic group. Participant characteristics are summarized in Table 1.

**Table 1.** Participant characteristics.

| Characteristic | Overall (n=27) | Menstruating<br>(n=13) | Amenorrheic<br>(n=14) |
| --- | --- | --- | --- |
| Age, years | 51.0<br>[49.0-53.5] | 49.0 [48.0-50.0] | 53.5 [53.0-54.0] |
| BMI, kg/m<sup>2</sup> | 21.0 [19.4-22.8] | 20.5 [18.9-22.1] | 21.5 [20.2-23.1] |
| FSH, mIU/mL | 44.8 [10.3-70.2] | 8.5 [5.8-19.4] | 70.2 [53.5-79.9] |
| E2, pg/mL | 45.6 [5.0-105.5] | 107 [58.3-222] | 5.0 [5.0-6.3] |
| E2 <5.0 pg/mL, n/N | 10/27 | 0/13 | 10/14 |
| Frequency-adjusted<br>symptom burden<br>score, 0-150 | 45.0 [6.5-58.5] | 36.0 [0-51.0] | 45.0 [22.5-60.8] |
| Vasomotor score, 0-30 | 6.0 [0-16.5] | 3.0 [0-16.0] | 12.5 [5.3-19.0] |
| Psychological score,<br>0-45 | 12.0 [1.0-19.0] | 6.0 [0-19.0] | 14.0 [4.5-19.0] |

Values are median [interquartile range] unless otherwise indicated. BMI, body mass index; E2, estradiol; FSH, follicle-stimulating hormone. For descriptive calculations, E2 values below 5.0 pg/mL were provisionally assigned a value of 5.0 pg/mL. The frequency-adjusted symptom burden score was calculated as the sum of severity × frequency products across the 10 symptom items.

The median overall frequency-adjusted symptom burden score was 45.0 (IQR, 6.5-58.5). The median vasomotor symptom score was 6.0 (IQR, 0-16.5), and the median psychological symptom score was 12.0 (IQR, 1.0-19.0). The score distributions included multiple zero scores and tied values, supporting the use of rank-based correlation.

**3.2 Overall frequency-adjusted symptom burden: correlations across all protein spots**

Analysis of the serum 2-DE images using SameSpots identified 550 matched protein spots (Fig. 1B). Spot intensities were normalized to the total intensity of all spots in each gel. Spearman rank correlations were calculated between the normalized volume of each spot and the frequency-adjusted symptom burden score, and P values from the 550 tests were adjusted using the Benjamini-Hochberg FDR procedure (Table S1). No spot remained significant after FDR adjustment (minimum q = 0.1173), although several spots showed strong nominal associations (minimum unadjusted P = 0.0004). Thirteen spots met the screening criteria of |ρ| ≥ 0.50 and nominal P < 0.01 (Table 2). These spots were qualitatively reviewed for spot morphology, signal intensity, separation from neighboring spots, and matching consistency across gels. Based on the combination of statistical screening and image characteristics, spots #285 and #636 were prioritized for subsequent domain-specific analyses. The frequency-adjusted symptom burden score was inversely correlated with spot #285 (ρ = -0.6150; 95% CI, -0.8106 to -0.2960; nominal P = 0.0006; FDR q = 0.1173; Fig. 2) and spot #636 (ρ = -0.5127; 95% CI, -0.7523 to -0.1533; nominal P = 0.0062; FDR q = 0.2644; Fig. 3).

**Fig. 2.**
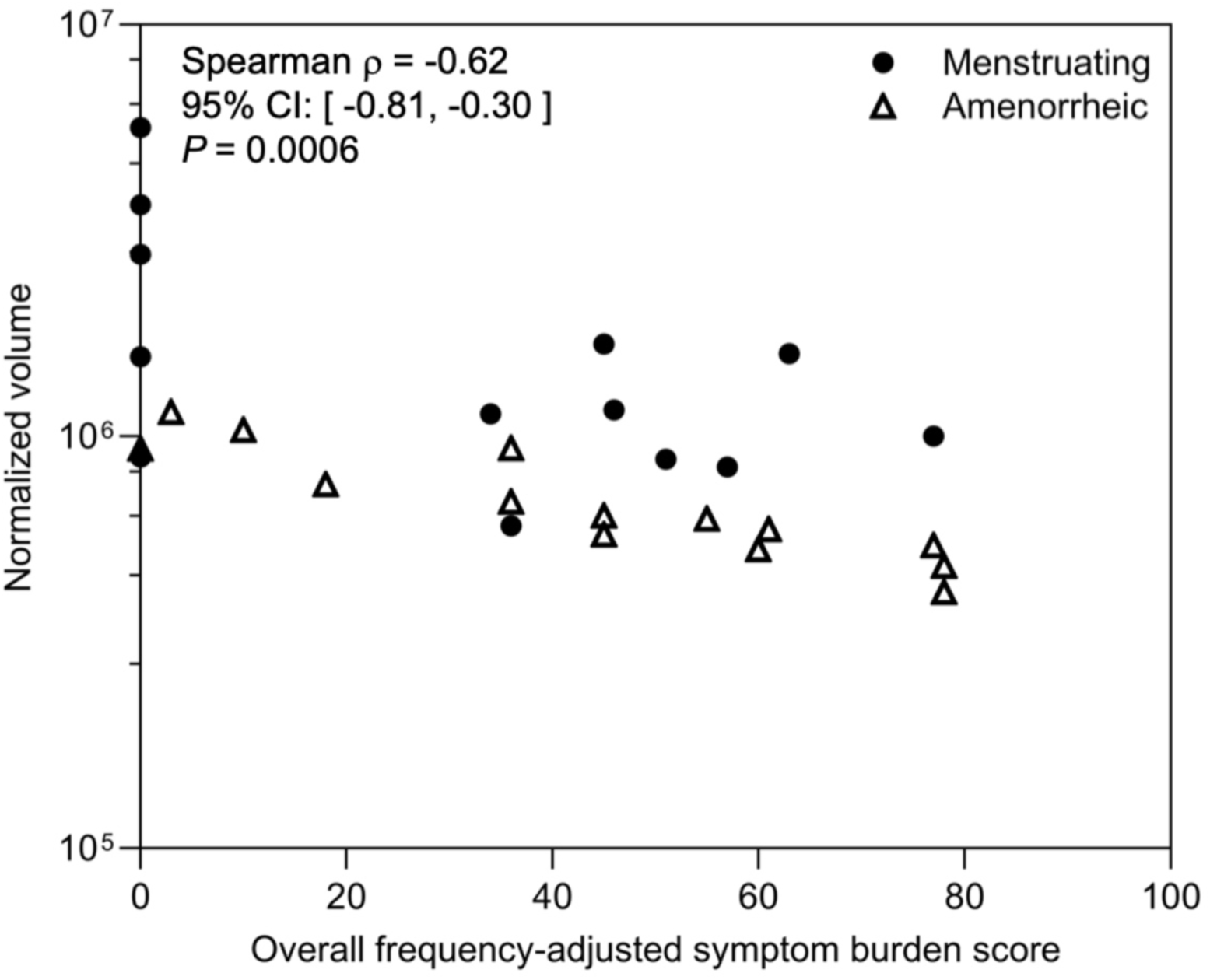
Association between spot #285 normalized volume and overall frequency-adjusted symptom burden score. The frequency-adjusted symptom burden score was calculated as the sum of severity × frequency products across 10 menopausal symptom items. Filled circles indicate menstruating participants and open triangles indicate amenorrheic participants. The y-axis is logarithmic. The Spearman ρ, 95% confidence interval (CI), and P value shown in the figure refer to the overall two-sided Spearman rank correlation.

**Fig. 3.**
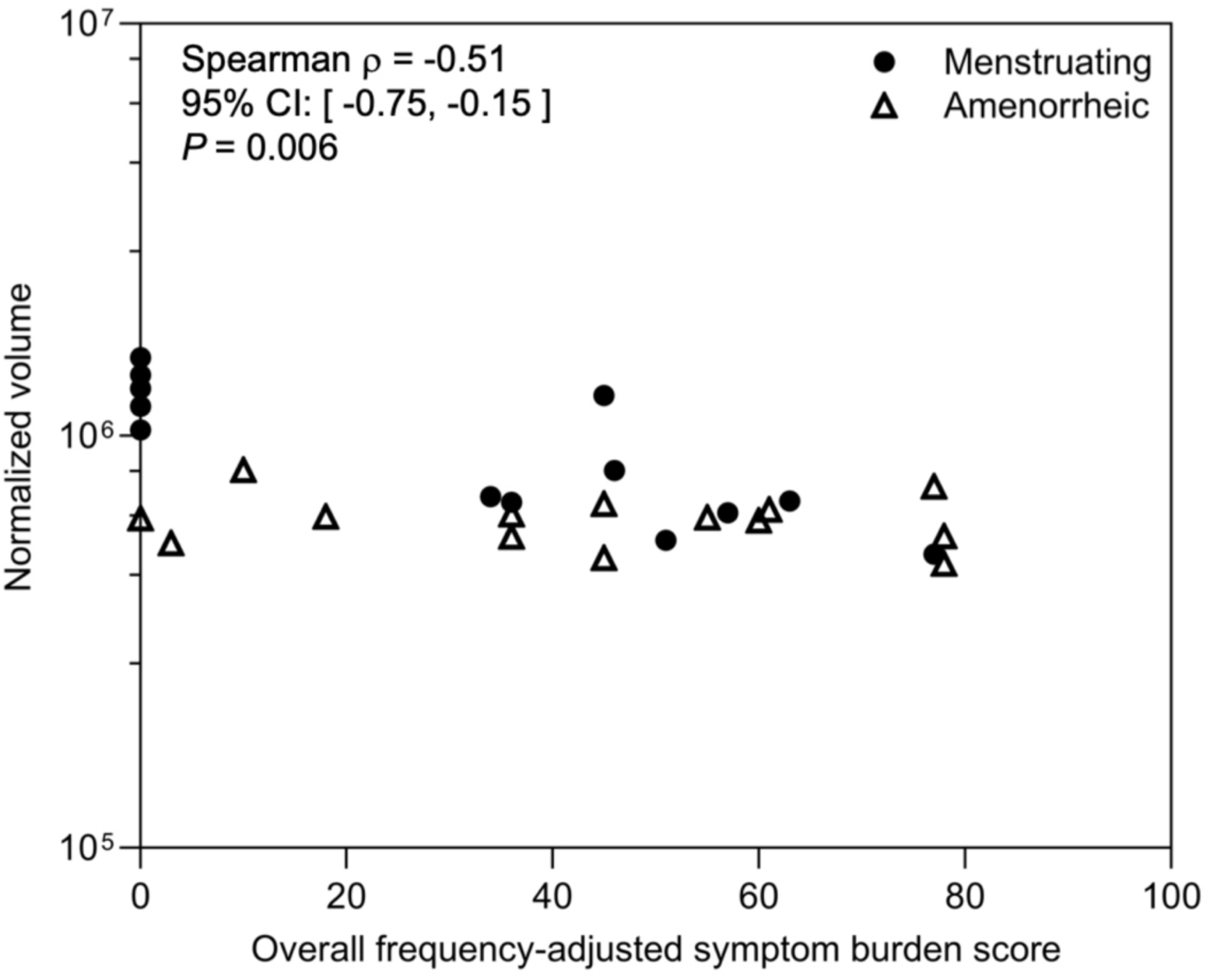
Association between spot #636 normalized volume and overall frequency-adjusted symptom burden score. The frequency-adjusted symptom burden score was calculated as the sum of severity × frequency products across 10 menopausal symptom items. Filled circles indicate menstruating participants and open triangles indicate amenorrheic participants. The y-axis is logarithmic. The Spearman ρ, 95% CI, and P value shown in the figure refer to the overall two-sided Spearman rank correlation.

**Table 2.** Candidate protein spots identified in the 550-spot screen of the frequency- adjusted symptom burden score.

| Spot | Spearman's $\rho$ | P | FDR q |
| --- | --- | --- | --- |
| 150 | 0.6347 | 0.0004 | 0.1173 |
| 538 | 0.6157 | 0.0006 | 0.1173 |
| 285 | -0.6150 | 0.0006 | 0.1173 |
| 283 | -0.5748 | 0.0017 | 0.2356 |
| 266 | -0.5490 | 0.0030 | 0.2644 |
| 286 | -0.5459 | 0.0032 | 0.2644 |
| 257 | -0.5410 | 0.0036 | 0.2644 |
| 450 | -0.5302 | 0.0044 | 0.2644 |
| 267 | -0.5247 | 0.0050 | 0.2644 |
| 155 | 0.5225 | 0.0052 | 0.2644 |
| 779 | -0.5182 | 0.0056 | 0.2644 |
| 844 | -0.5151 | 0.0060 | 0.2644 |
| 636 | -0.5127 | 0.0062 | 0.2644 |

The frequency-adjusted symptom burden score was calculated as the sum of severity × frequency products across the 10 symptom items. Nominal P values are shown together with Benjamini-Hochberg FDR q values.

### 3.3 Spot #285 and vasomotor symptoms

The vasomotor symptom score was the sum of the hot-flush and sweating scores (possible range, 0-30). Spot #285 normalized volume was inversely associated with vasomotor symptom score in the overall cohort (Spearman ρ = -0.6697; 95% CI, -0.8403 to -0.3785; P = 0.0001; Fig. 4). Thus, higher vasomotor symptom scores were monotonically associated with lower spot #285 normalized volumes.

**Fig. 4.**
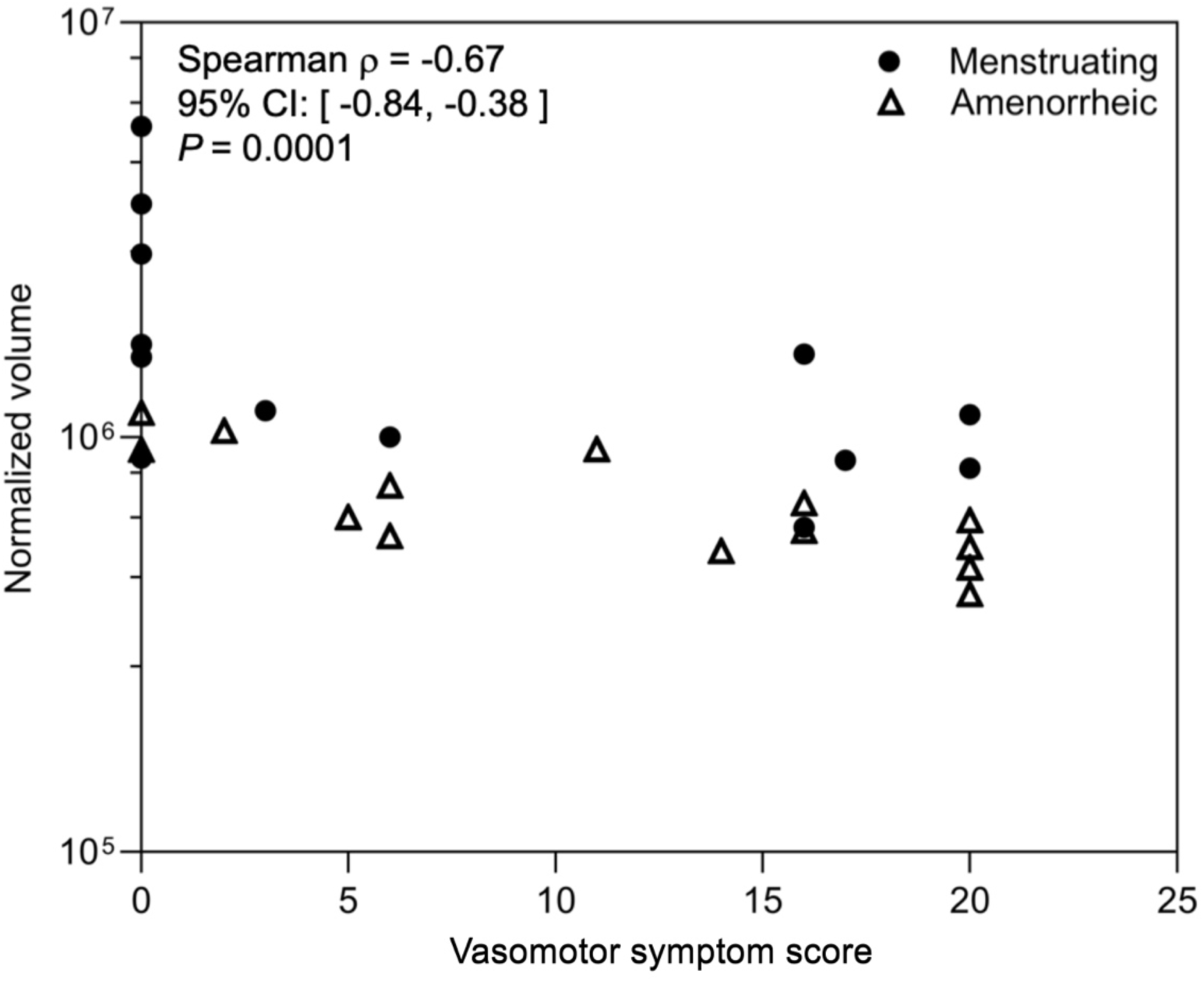
Association between spot #285 normalized volume and vasomotor symptom score. The vasomotor symptom score was calculated as the sum of the severity × frequency scores for hot flushes and sweating (possible range, 0–30). Filled circles indicate menstruating participants and open triangles indicate amenorrheic participants. The y-axis is logarithmic. The Spearman ρ, 95% CI, and P value shown in the figure refer to the overall two-sided Spearman rank correlation.

### 3.4 Spot #636 and psychological symptoms

The psychological symptom score was the sum of the depressed mood, irritability, and sleep disturbance scores (possible range, 0-45). Spot #636 normalized volume was inversely associated with psychological symptom score in the overall cohort (Spearman ρ = -0.4824; 95% CI, -0.7343 to -0.1138; P = 0.0108; Fig. 5). Thus, higher psychological symptom scores were associated with lower spot #636 normalized volumes.

**Fig. 5.**
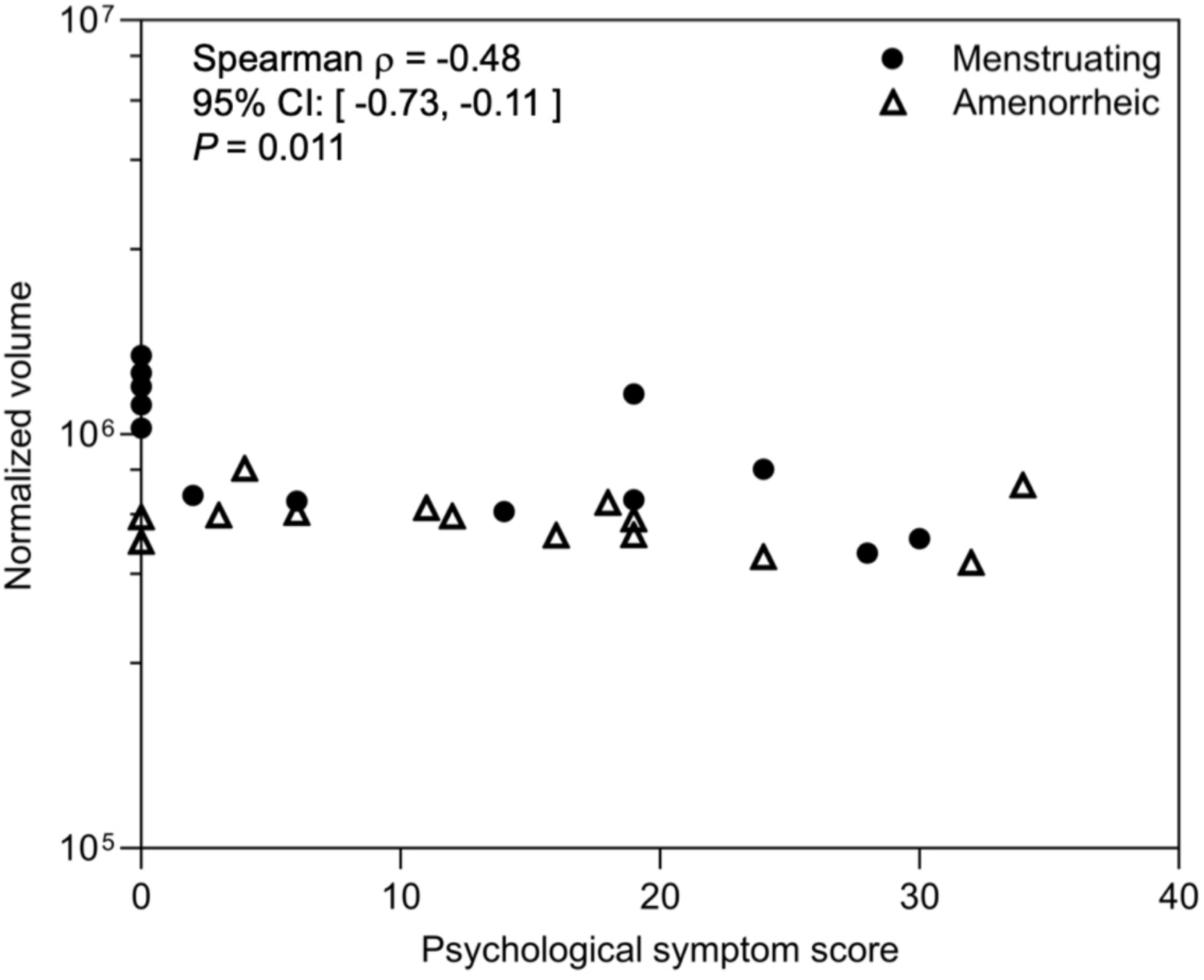
Association between spot #636 normalized volume and psychological symptom score. The psychological symptom score was calculated as the sum of the severity × frequency scores for sleep disturbance, irritability, and depressed mood (possible range, 0–45). Filled circles indicate menstruating participants and open triangles indicate amenorrheic participants. The y-axis is logarithmic. The Spearman ρ, 95% CI, and P value shown in the figure refer to the overall two-sided Spearman rank correlation.

### 3.5 Correlations separately in menstruating and amenorrheic participants

For spot #285, inverse correlations with vasomotor symptom score were observed among menstruating participants (ρ = -0.6783; 95% CI, -0.8984 to -0.1856; P = 0.0138) and amenorrheic participants (ρ = -0.7626; 95% CI, -0.9233 to -0.3748; P = 0.0023) (Table 3). For spot #636, the inverse correlation with psychological symptom score was strong among menstruating participants (ρ = -0.7500; 95% CI, -0.9233 to -0.3228; P = 0.0047) but was not evident among amenorrheic participants (ρ = -0.1894; 95% CI, -0.6641 to 0.3941; P = 0.5135) (Table 3). The stratified estimates therefore differed in magnitude for spot #636, with a stronger inverse correlation among menstruating participants. Given the small subgroup sizes, the post hoc nature of the stratified analysis, and the absence of a formal between-group comparison, this pattern should be interpreted descriptively and requires independent confirmation.

**Table 3.** Spearman rank correlations between symptom scores and normalized spot volumes, stratified by menstrual status.

| Spot | Group | Spearman's $\rho$ | 95% CI | P |
| --- | --- | --- | --- | --- |
| 285 | Menstruating | -0.6783 | -0.8984, -0.1856 | 0.0138 |
| 285 | Amenorrheic | -0.7626 | -0.9233, -0.3748 | 0.0023 |
| 636 | Menstruating | -0.7500 | -0.9233, -0.3228 | 0.0047 |
| 636 | Amenorrheic | -0.1894 | -0.6641, 0.3941 | 0.5135 |

Spearman correlations between spot #285 normalized volume and vasomotor symptom score, and between spot #636 normalized volume and psychological symptom score, estimated separately in menstruating and amenorrheic participants. P values are nominal and were not adjusted for the candidate-selection process.

## 4. Discussion

In this exploratory study, a complete 550-spot screen followed by qualitative image review prioritized serum 2-DE spots #285 and #636 for focused analysis. The full-spot screen is important for interpreting the candidate-focused results. Thirteen spots met the effect-size and nominal-P screening criteria, but none remained significant after FDR adjustment.

Accordingly, spots #285 and #636 were not selected solely because they represented the statistically strongest signals. Rather, they were prioritized through a combined process that considered the statistical screening and whether the corresponding 2-DE features were sufficiently well defined and technically suitable for subsequent isolation, identification, and assay development. This pragmatic approach is relevant to exploratory proteomic discovery, but the qualitative image review introduces additional selection flexibility and should therefore be interpreted transparently.

The frequency-adjusted symptom burden score was inversely associated with both spots #285 and #636. Given the small sample size and the large number of spot-wise tests, statistical power after multiplicity adjustment was limited; these findings should therefore be interpreted as exploratory. Spot #285 showed the most coherent pattern across analysis levels. Higher overall frequency-adjusted symptom burden and higher vasomotor symptom scores were both associated with lower spot #285 normalized volume. In addition, inverse associations with vasomotor symptom score were observed in both menstruating and amenorrheic participants. Although the subgroup estimates were similar in direction, the small subgroup sizes preclude firm conclusions regarding equivalence between menstrual-status groups.

Spot #636 was inversely associated with both overall frequency-adjusted symptom burden and the psychological symptom composite. However, the menstrual-status-stratified estimates differed in magnitude: a strong inverse correlation was observed among menstruating participants, whereas no clear association was observed among amenorrheic participants. No formal between-group comparison was performed, so this pattern should not be interpreted as evidence of a statistically established difference between menstrual-status groups. The observed pattern could reflect biological differences related to reproductive or endocrine status, confounding by other participant characteristics, or instability arising from the small subgroup sizes. It would therefore be premature to describe spot #636 as a uniform marker of psychological symptoms across menstrual states. Proteomic research specifically addressing psychological symptoms during the menopausal transition remains limited, and further studies are needed to determine whether spot #636 represents a reproducible biological correlate of psychological symptom burden.

This study has several limitations. First, the sample size was small, and no independent validation cohort was available. Second, the cohort included both menstruating and amenorrheic women, and the analyses did not adjust for potential confounders such as age, E2, FSH, body mass index, hormone therapy, medication use, comorbidities, or menstrual- cycle timing. Third, the qualitative image review formed part of candidate prioritization and therefore introduced an additional exploratory selection step. Finally, biological interpretation is currently limited because spots #285 and #636 have not been molecularly identified. Individual 2-DE spots may represent intact proteins, fragments, modified proteoforms, or co- migrating species.

A follow-up study should prespecify the candidate spots and scoring rules, include an independent validation cohort, document technical precision and batch effects, and assess associations using multivariable models. Diagnostic or monitoring performance should be evaluated using receiver operating characteristic (ROC) analysis, calibration, and clinically meaningful reference thresholds only after the molecular identities of the candidate spots have been established and suitable analytical assays developed. Molecular identification should include spot excision followed by LC-MS/MS, confirmation of the apparent molecular mass and isoelectric point, and orthogonal measurement using antibody-based methods or targeted mass spectrometry.

## Data Availability

The anonymized datasets generated and/or analyzed during the current study are available from the corresponding author on reasonable request, subject to ethical and institutional approval where applicable.

## Acknowledgments

The authors thank Hiroyuki Mabuchi, President and Chief Executive Officer of aiwell Inc., for institutional support and for facilitating the implementation of this research project. The authors also thank Kaoru Nagase, Hayato Nakamura and Taro Mori of aiwell Inc. for their administrative support and assistance with project coordination.

## Author Contributions

Maki Sasanuma: Methodology, Investigation, Formal analysis, Visualization, Writing – original draft, and Project administration. Misa Kuroki: Formal analysis and Writing – review and editing. Hirotaka Tabata: Investigation and Data curation. Atsushi Kajiwara: Investigation and Data curation. Akiko Shiraki: Investigation and Data curation. Rehab F. Abdelhamid: Investigation and Data curation. Yukoh Nakazaki: Conceptualization, Formal analysis, Supervision, Funding acquisition, and Writing – review and editing. Miho Takao: Conceptualization, Investigation, and Resources. All authors reviewed and approved the final manuscript.

